# Protection against symptomatic SARS-CoV-2 BA.5 infection conferred by the Pfizer-BioNTech Original/BA.4-5 bivalent vaccine compared to the mRNA Original (ancestral) monovalent vaccines – a matched cohort study in France

**DOI:** 10.1101/2023.03.17.23287411

**Authors:** Vincent Auvigne, Cynthia Tamandjou, Justine Schaeffer, Sophie Vaux, Isabelle Parent du Chatelet

**Affiliations:** Direction des maladies infectieuses - Unité infections respiratoires et vaccination, Santé publique France, Public Health France. F-94415 Saint-Maurice, France

## Abstract

This cohort study aimed to evaluate the protection against symptomatic SARS-CoV-2 infection conferred by the Pfizer-BioNTech Original/BA.4-5 bivalent vaccine compared to mRNA Original (ancestral) monovalent vaccines. Individuals of ≥60 years old who received a booster dose between 03/10/2022 and 06/11/2022, when both the bivalent and monovalent vaccines were used in France, were included. Individuals who received a booster dose with (1) a monovalent Original mRNA vaccine (Pfizer- BioNTech or Moderna) or (2) the bivalent Pfizer-BioNTech Original/BA.4-5 vaccine were matched. The outcome of interest was a positive SARS-CoV-2 RT-PCR or antigenic test associated to self-reported symptoms, at least seven days after receiving the booster dose. Data were analysed with a Cox Proportional-Hazards model adjusted for the presence of previous infection, age, sex, and the presence of medium risk comorbidities. A total of 136,852 individuals were included and followed for a median period of 77 days. The bivalent vaccine conferred an additional protection of 8% [95% CI: 0% - 16%, p=0.045] against symptomatic SARS-CoV-2 infection compared to the monovalent vaccines.

## Introduction

Since the beginning of the COVID-19 pandemic, the evolution of SARS-CoV-2 and vaccine- and infection-induced selection pressure has led to increasing viral immune escape. Consequently, vaccines targeted towards the original SARS-CoV-2 strain showed reduced effectiveness against Omicron infection. The flexibility of the mRNA technology has allowed the rapid design of new vaccines, adapted to Omicron sublineages BA.1 and BA.4/5, henceforth referred as bivalent vaccines. In France, bivalent vaccines were used from early October 2022. *In vitro* studies evaluating antibody levels following bivalent booster vaccination and their ability to neutralize more recent variants have shown conflicting results, with increased or similar neutralisation of BA.4, BA.5 and some of their sublineages following a booster with a bivalent vaccine compared to an original monovalent vaccine.^1,2^ To date, real-world studies have compared individuals who have recently received a bivalent booster to individuals without recent booster.^3,4^ The contribution of these bivalent vaccines compared to the original vaccines is therefore questionable. This study aimed to evaluate the protection against symptomatic SARS-CoV-2 infection conferred by the Pfizer-BioNTech Original/BA.4-5 bivalent vaccine compared to the mRNA Original (ancestral) monovalent vaccines.

## Methods

We built a retrospective matched closed cohort using data from the French national surveillance databases on COVID-19 vaccination (VAC-SI) and virological tests (SIDEP). The cohort included 60 years and older individuals who received a booster dose between 03/10/2022 and 06/11/2022, when both bivalent and monovalent vaccines were used in France. Individuals with very-high-risk comorbidities and those who received a primary vaccination schedule other than two doses were excluded (Supplementary appendix, Table S1). Two arms of the cohort were formed: individuals who received a booster dose with (1) the monovalent Original mRNA vaccine (Pfizer-BioNTech or Moderna) or (2) the bivalent Pfizer-BioNTech Original/BA.4-5 vaccine. The outcome of interest was a positive COVID-19 virological diagnosis (by RT-PCR or antigenic test) associated to self-reported symptoms, at least seven days after receiving the booster dose. The follow-up period was from 10/10/2022 to 06/03/2023; then, the dominant variants were BA.5 sublineages and particularly BQ.1.1.

Participants from both arms were matched to minimise bias resulting from differences in exposure to the risk of infection between the two arms of the study. The matching variables included the booster rank (inclusion at 1st, 2nd or 3rd booster dose), the week of booster vaccination, and the area of residence (Table S2). The booster rank was considered for the matching as it might be related to age and possibly to risk factors and behaviours. The week of booster vaccination allowed to take into account the evolution of the risk of infection over time, as incidence rate in France in the over 60s varied from 300 to 700/100,000 over the study follow-up period (Figure S1). The area of residence corrected for geographical differences in term of incidence and variant circulation; a replacement of other BA.5 sublineages by the BQ.1.1 sublineage was observed over the follow-up period, with an earlier spread in the Ile-de-France region (Figure S2). Multivariate analysis was performed using a Cox Proportional-Hazards Model, adjusted for the presence of previous infection, age, sex, and the presence of medium risk comorbidities. Medium-risk comorbidities included obesity, diabetes, chronic renal failure, chronic obstructive pulmonary disease, respiratory failure, hypertension, heart failure.

Previous infection was classified according to the dominant variants in France at the time of infection: Delta/Pre-Delta, Transition from Delta to BA.1, Omicron BA.1 (including the transition between BA.1 and BA.2) and Omicron BA.2 BA.4/5. For individuals with multiple documented previous infections, the most recent was selected. A sensitivity analysis limited to individuals with no history of infection prior to the inclusion was performed. Results of the Cox model, the adjusted hazard ratios (aHR), are presented as percent of additional protection using the formula: Additional protection (%) = (1 – aHR) * 100.

## Results

A total of 136,852 individuals were included in the cohort. Eighty-five percent of these individuals were included following their second booster (Table 1, Table 2, Figure S3). The vaccines of the inclusion boosters in the Original arm were Pfizer Original (96%) and Moderna Original (4%). During a median follow-up period of 77 days, we observed 1,147 positive SARS-CoV-2 virological tests in the Original arm (individuals who received a booster dose with the mRNA vaccines targeting the original SARS-CoV-2 strain) and 1,025 in the Original/BA.4-5 arm (individuals who received a booster dose with the bivalent Original/BA.4-5 Pfizer-BioNTech vaccine) (Table 1).

**Table 1:** Descriptive statistics by inclusion booster

|  | Inclusion booster |  | p-value<sup>2</sup> |
| --- | --- | --- | --- |
|  | Original, N = 68,426<sup>1</sup> | Original / BA.4-5, N = 68,426<sup>1</sup> |  |
| Week of inclusion |  |  | >0.99 |
| 2022-W40 | 10,244 (15%) | 10,244 (15%) |  |
| 2022-W41 | 26,285 (38%) | 26,285 (38%) |  |
| 2022-W42 | 16,937 (25%) | 16,937 (25%) |  |
| 2022-W43 | 10,087 (15%) | 10,087 (15%) |  |
| 2022-W44 | 4,873 (7%) | 4,873 (7%) |  |
| Booster dose at inclusion |  |  | >0.99 |
| 1 | 1,586 (2%) | 1,586 (2%) |  |
| 2 | 58,180 (85%) | 58,180 (85%) |  |
| 3 | 8,660 (13%) | 8,660 (13%) |  |
| Sex |  |  | 0.021 |
| Female | 37,535 (55%) | 37,109 (54%) |  |
| Male | 30,891 (45%) | 31,317 (46%) |  |
| Age |  |  | <0.001 |
| 60-79 | 49,355 (72%) | 51,563 (75%) |  |
| 80-99 | 19,071 (28%) | 16,863 (25%) |  |
| Previous infection |  |  | <0.001 |
| Undocumented | 47,870 (70%) | 46,047 (67%) |  |
| Delta/Pre-Delta | 1,355 (2%) | 1,384 (2%) |  |
| Tr_Delta-BA.1 | 842 (1%) | 894 (1%) |  |
| Omicron BA.1 | 5,866 (9%) | 6,217 (9%) |  |
| Omicron BA.2 BA.4/5 | 12,493 (18%) | 13,884 (20%) |  |
| Medium-risk comorbidity |  |  | <0.001 |
| No | 42,636 (62%) | 44,936 (66%) |  |
| Yes | 25,790 (38%) | 23,490 (34%) |  |
| Outcome: All positive symptomatic tests |  |  | 0.008 |
| 0 | 67,279 (98%) | 67,401 (99%) |  |
| 1 | 1,147 (2%) | 1,025 (1%) |  |
| Vaccination post inclusion (censoring) |  |  | <0.001 |
| No | 68,305 (100%) | 68,378 (100%) |  |
| Yes | 121 (0%) | 48 (0%) |  |

<sup>1</sup>n (%)
<sup>2</sup>Pearson's Chi-squared test

**Table 2:** Immunisation history by inclusion booster

|  | Inclusion booster |  | p-value<sup>2</sup> |
| --- | --- | --- | --- |
|  | Original, N = 68,426<sup>1</sup> | Original / BA.4-5, N = 68,426<sup>1</sup> |  |
| Vaccine 1 |  |  |  |
| Pfizer Original | 47,051 (69%) | 47,724 (70%) |  |
| Moderna Original | 5,545 (8%) | 4,941 (7%) |  |
| AstraZeneca | 15,826 (23%) | 15,757 (23%) |  |
| Janssen | 3 (0%) | 3 (0%) |  |
| Pfizer paed. | 1 (0%) | 0 (0%) |  |
| Novavax | 0 (0%) | 1 (0%) |  |
| Vaccine 2 |  |  |  |
| Pfizer Original | 47,621 (70%) | 48,465 (71%) |  |
| Moderna Original | 5,844 (9%) | 5,251 (8%) |  |
| AstraZeneca | 14,953 (22%) | 14,707 (21%) |  |
| Janssen | 6 (0%) | 2 (0%) |  |
| Pfizer paed. | 2 (0%) | 0 (0%) |  |
| Novavax | 0 (0%) | 1 (0%) |  |
| Vaccine 3 (inclusion booster 1 excluded) |  |  | <0.001 |
| Pfizer Original | 54,374 (81%) | 56,136 (84%) |  |
| Moderna Original | 12,466 (19%) | 10,704 (16%) |  |
| NA | 1,586 | 1,586 |  |
| Vaccine 4 (inclusion booster 2 excluded) |  |  | <0.001 |
| Pfizer Original | 7,991 (92%) | 8,162 (94%) |  |
| Moderna Original | 669 (8%) | 498 (6%) |  |
| NA | 59,766 | 59,766 |  |

<sup>1</sup>n (%)
<sup>2</sup>Pearson's Chi-squared test

The additional protection against symptomatic SARS-CoV-2 infection with the bivalent vaccine compared to original vaccines was statistically significant but minimal: 8% [95% CI: 0% - 16%; p=0·045]. Previous infection conferred additional protection; the most recent infections related to variants close to those circulating during the follow-up period were the most protective. For instance, a previous infection with BA.2 or BA.4-5 conferred an additional protection of 74% [95% CI: 78% - 69%] (Figure 1, Table S4). Older previous infections, caused by variants more distinct from the circulating strains, conferred lower but still detectable protection in boosted participants. The analysis restricted to participants without documented previous infection showed no significant difference (Table S5).

**Figure 1:**
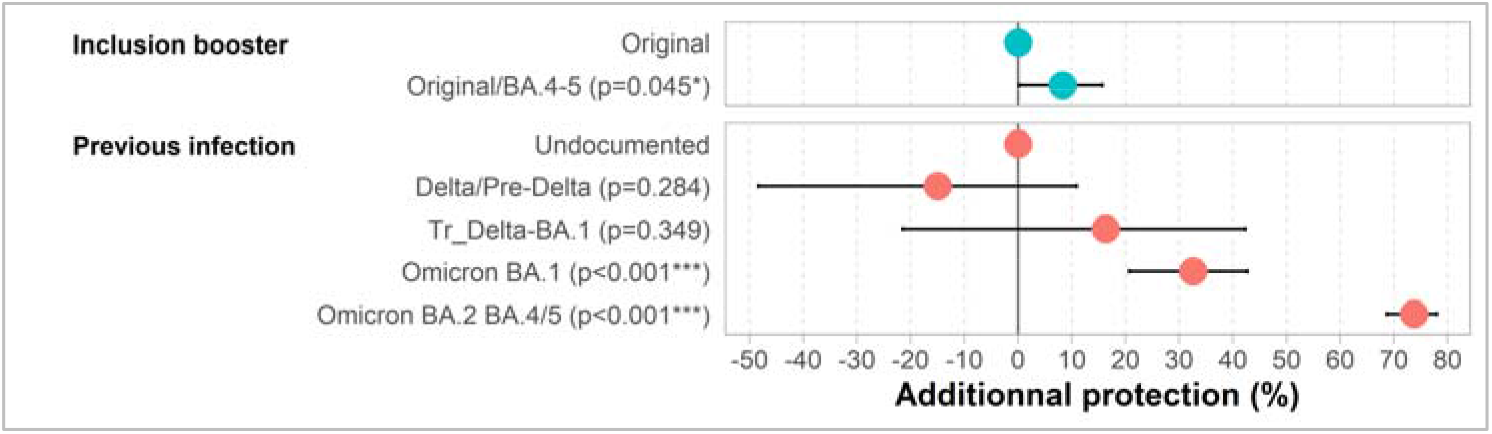
Adjusted additional protection against symptomatic SARS-CoV-2 infection of a booster with Pfizer-BioNTech Original/BA.4-5 bivalent vaccine compared to a booster with mRNA Original vaccines, and of previous infection. Multivariate Cox proportional-hazards model adjusted on age, sex and presence of medium-risk comorbidities-N=136,852 individuals ≥ 60 years old without high-risk-comorbidities. During a median follow-up time of 77 days of BA2 and BA.4/5 dominance, 2,172 infections were observed.

## Conclusion

The studied bivalent vaccine only partially compensated the immune evasion ability of the Omicron variant and its sublineages. Still, it is important to bear in mind that a recent booster whether with an original monovalent or a bivalent vaccine offers additional protection against symptomatic SARS-CoV-2 infection compared to an outdated booster dose.^3–5^ Therefore, booster vaccination remains important to maximize protection against COVID-19, especially for the most vulnerable populations.

## Data Availability

While all data used in this analysis were pseudonymised, the individual-level nature of the data used risks individuals being identified, or being able to self-identify, if it is released publicly. Requests for access to the underlying source data should be directed to Sante publique France and will be granted in accordance with the GDPR and French Law.

## Ethical statement

The Data Protection Officer of Santé Publique France specified that the opinion of an ethics committee was not required.

## Acknowledgments

The authors thank Daniel Levy-Bruhl, Yann Le Strat and Catarina Krug for their methodological support, Charline Montagnat for her support in data management and Bruno Coignard and Didier Che for their valuable support in the implementation and the realization of this study

