## Supplementary material for "Protection against symptomatic SARS-CoV-2 BA.5 infection conferred by the Pfizer-BioNTech Original/BA.4-5 bivalent vaccine compared to the mRNA Original (ancestral) monovalent vaccines – a matched cohort study in France"

### Appendix

#### Selection of individuals

*Table S1 : Inclusion process (before matching)*

| | Excluded | Remain |
|---|---|---|
| Born in 1962 and earlier and boostered during the inclusion period | | 878,492 |
| Exclusion for data inconsistency | 10,354 | 868,138 |
| very-high-risk comorbidity[1] | 91,686 | 773,623 |
| primary vaccine series with less or more than two doses | 47,565 | 728,887 |
| at least one supplementary dose for medical reason (ex: immunosuppression) | 24,889 | 703,998 |
| pre-inclusion booster with non mRNA vaccine | 113 | 703,885 |
| positive test < 60 days before follow-up | 1,861 | 702,024 |
| >20 tests in the 6 months prior to inclusion | 26 | 701,998 |
| over 99 years | 1,940 | **700,058** |

[1] : cancers, haematological malignancies undergoing chemotherapy, severe chronic kidney disease, chronic dialysis, solid organ transplants, haematopoietic stem cell allografts, chronic multi-disease conditions with two or more organ failures, certain rare diseases and those at particular risk of infection, and Down's syndrome

*Table S2 : Size of the study population after matching*

| | Total | matched | % matched |
|---|---|---|---|
| Original/BA.4-5 | 582,398 | 68,426 | 12% |
| Original | 117,660 | 68,426 | 58% |
| Total | 697,649 | 136,852 | 20% |

#### Epidemiological background

*Figure S1: Weekly community incidence rates (France, data SI-DEP)*

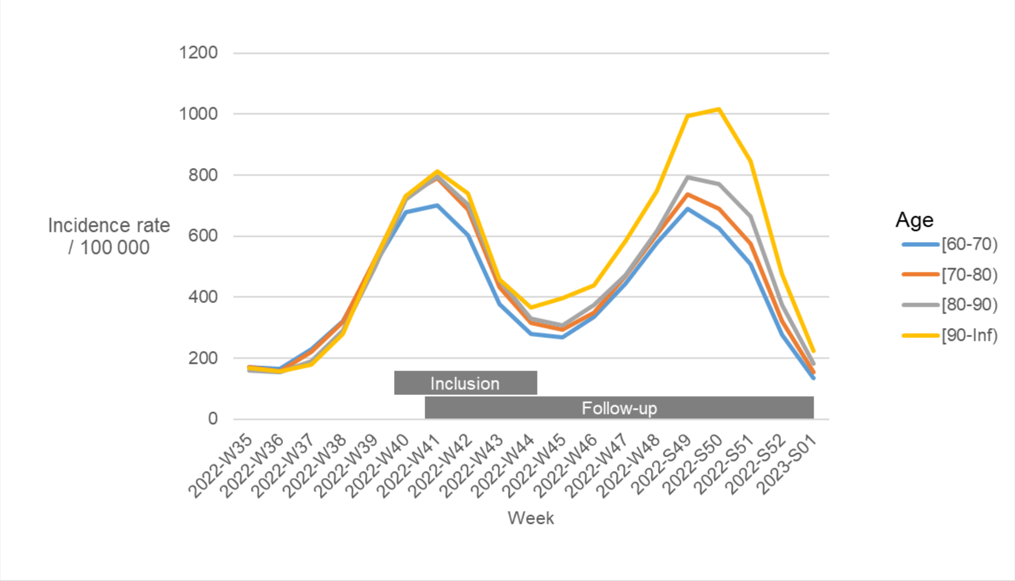

*Figure S2: Emergence of B.Q.1.1* in France according to representative whole genome sequencing*

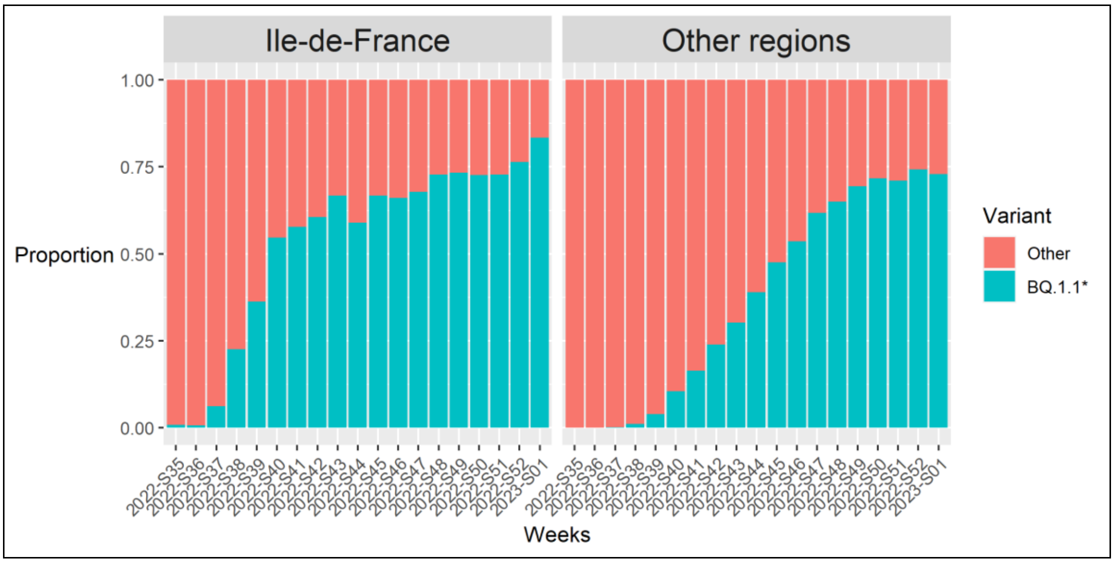

#### Descriptive and univariate analysis

*Figure S3 : Immunisation history by rank of booster at inclusion (one colour per dose: salmon: dose 1, pink: dose 2, blue: dose 3, light blue: dose 4, green: dose 5)*

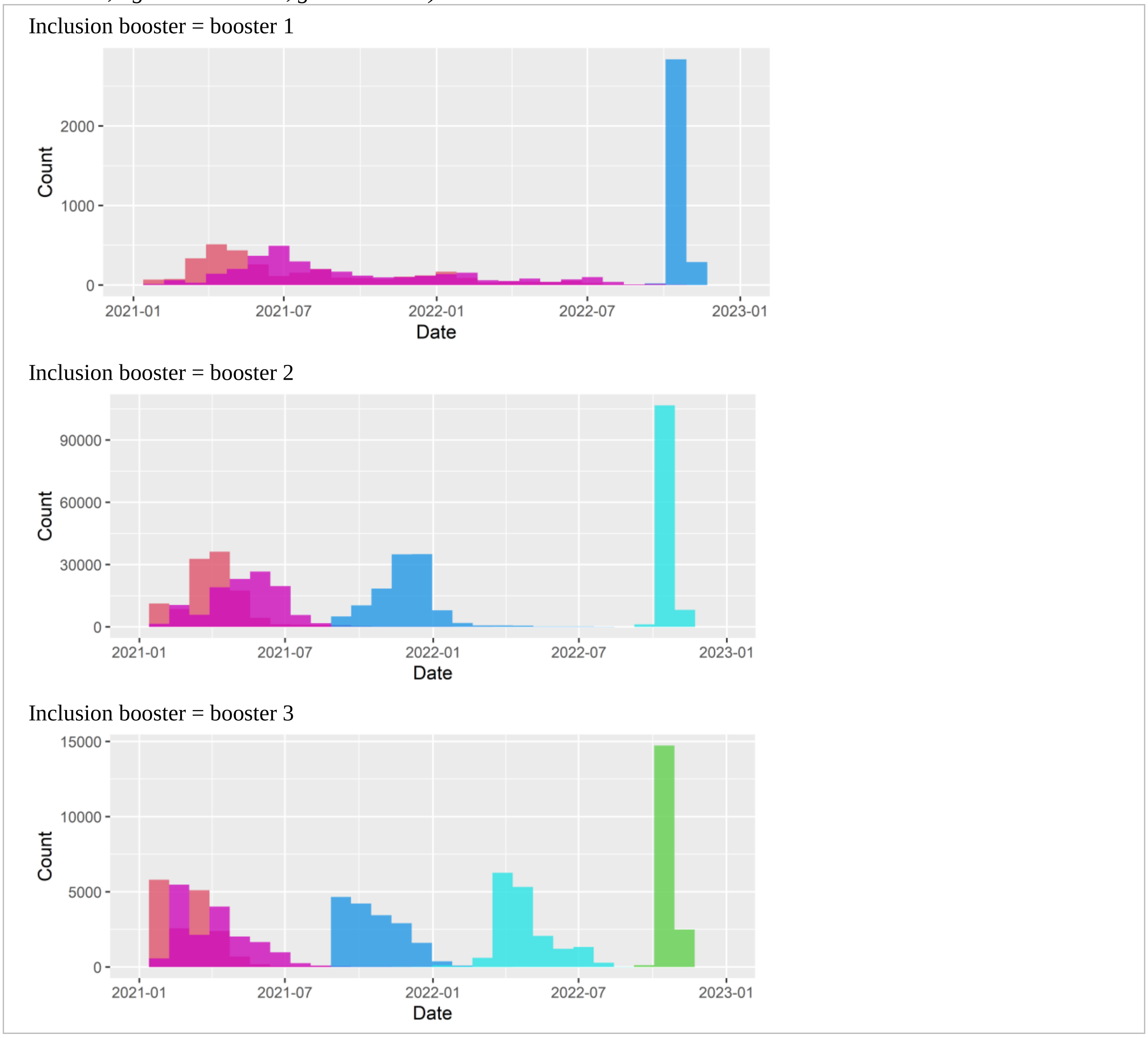

#### Univariate analysis

*Tableau S3 : Descriptive statistics by occurrence of the outcome*

| | Outcome | | |
|---|---|---|---|
| | No, N = 134,680[1] | Yes, N = 2,172[1] | p-value[2] |
| Week of inclusion | | | <0.001 |
| 2022-W40 | 20,097 (98%) | 391 (2%) | |
| 2022-W41 | 51,672 (98%) | 898 (2%) | |
| 2022-W42 | 33,367 (99%) | 507 (1%) | |
| 2022-W43 | 19,921 (99%) | 253 (1%) | |
| 2022-W44 | 9,623 (99%) | 123 (1%) | |
| Inclusion booster | | | 0.008 |
| Original | 67,279 (98%) | 1,147 (2%) | |
| Original / BA.4-5 | 67,401 (99%) | 1,025 (1%) | |
| Booster dose at inclusion | | | <0.001 |
| 1 | 3,126 (99%) | 46 (1%) | |
| 2 | 114,593 (98%) | 1,767 (2%) | |
| 3 | 16,961 (98%) | 359 (2%) | |
| Sex | | | <0.001 |
| Female | 73,552 (99%) | 1,092 (1%) | |
| Male | 61,128 (98%) | 1,080 (2%) | |
| Age | | | 0.89 |
| 60-79 | 99,319 (98%) | 1,599 (2%) | |
| 80-99 | 35,361 (98%) | 573 (2%) | |
| Previous infection | | | <0.001 |
| Undocumented | 92,126 (98%) | 1,791 (2%) | |
| Delta/Pre-Delta | 2,678 (98%) | 61 (2%) | |
| Tr_Delta-BA.1 | 1,708 (98%) | 28 (2%) | |
| Omicron BA.1 | 11,925 (99%) | 158 (1%) | |
| Omicron BA.2 BA.4/5 | 26,243 (99%) | 134 (1%) | |
| Medium-risk comorbidity | | | 0.003 |
| No | 86,249 (98%) | 1,323 (2%) | |
| Yes | 48,431 (98%) | 849 (2%) | |

[1]n (%)
[2]Pearson's Chi-squared test; Fisher's exact test

*Figure S4 : Cumulative probability over time of symptomatic infection since the booster dose by the type of vaccine of the booster dose - Follow-up started 7 days after booster injection; ribbons indicate 95%CI*

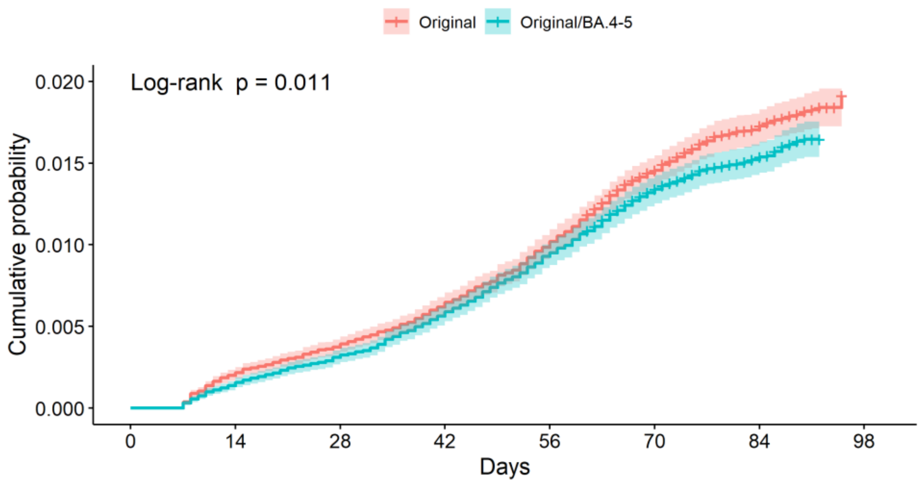

Numbers at risk (number of events)

| | 0 | 14 | 28 | 42 | 56 | 70 | 84 | 98 |
|---|---|---|---|---|---|---|---|---|
| Original | 68426 (0) | 68289 (148) | 68171 (268) | 68002 (443) | 67752 (698) | 61939 (989) | 34297 (1127) | 0 (1147) |
| Original/BA.4-5 | 68426 (0) | 68332 (108) | 68214 (223) | 68041 (403) | 67790 (650) | 62192 (909) | 34131 (1007) | 0 (1025) |

Days

Cumulative number of censoring

| | 0 | 14 | 28 | 42 | 56 | 70 | 84 | 98 |
|---|---|---|---|---|---|---|---|---|
| Original | 0 | 0 | 0 | 0 | 0 | 7469 | 38131 | 67279 |
| Original/BA.4-5 | 0 | 0 | 0 | 0 | 0 | 7446 | 40285 | 67401 |

Days

*Figure S5 : Cumulative probability over time of symptomatic infection since the booster dose by previous infection - Follow-up started 7 days after booster injection; ribbons indicate 95%CI*

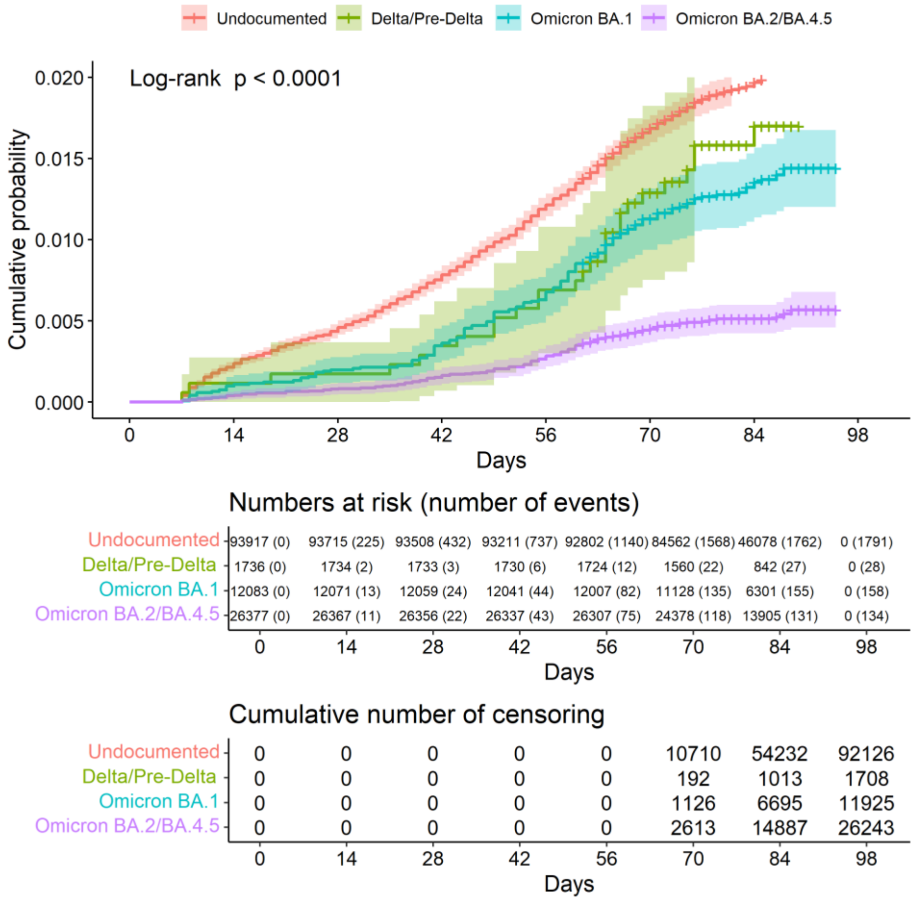

#### Multivariate analysis

*Table S4 : Multivariate analysis (Cox Proportional-Hazards Model - N=136,852, number of events= 2172)*

| | aHR[1] | 95% CI[1] | p-value |
|---|---|---|---|
| Inclusion booster | | | |
| Original | — | — | |
| Original / BA.4-5 | 0·92 | 0·84, 1·00 | 0·045 |
| Age | | | |
| 60-79 | — | — | |
| 80-99 | 0·98 | 0·89, 1·08 | 0·6 |
| Sex | | | |
| Female | — | — | |
| Male | 1·19 | 1·09, 1·29 | <0·001 |
| Medium-risk comorbidity | | | |
| No | — | — | |
| Yes | 1·12 | 1·03, 1·23 | 0·008 |
| Previous infection | | | |
| Undocumented | — | — | |
| Delta/Pre-Delta | 1·15 | 0·89, 1·48 | 0·3 |
| Tr_Delta-BA·1 | 0·84 | 0·58, 1·21 | 0·3 |
| Omicron BA.1 | 0·67 | 0·57, 0·79 | <0·001 |
| Omicron BA.2 BA.4/5 | 0·26 | 0·22, 0·31 | <0·001 |

[1]aHR = adjusted hazard ratio, CI = Confidence Interval

*Table S5 : Multivariate analysis – Participants without previous infection (Cox Proportional-Hazards Model - N=93,917, number of events= 1791)*

| | aHR[1] | 95% CI[1] | p-value |
|---|---|---|---|
| Inclusion booster | | | |
| Original | — | — | |
| Original / BA.4-5 | 0·96 | 0·88, 1·06 | 0·4 |
| Age | | | |
| 60-79 | — | — | |
| 80-99 | 0·90 | 0·81, 1·01 | 0·070 |
| Sex | | | |
| Female | — | — | |
| Male | 1·21 | 1·10, 1·33 | <0·001 |
| Medium-risk comorbidity | | | |
| No | — | — | |
| Yes | 1·12 | 1·02, 1·24 | 0·017 |

[1]aHR = Hazard Ratio, CI = Confidence Interval
